# Effects of a Ketogenic and Low Fat Diet on the Human Metabolome, Microbiome and Food-ome in Adults at Risk for Alzheimer’s Disease

**DOI:** 10.1101/2022.08.30.22279087

**Authors:** Amanda Hazel Dilmore, Cameron Martino, Bryan J. Neth, Kiana A. West, Jasmine Zemlin, Gibraan Rahman, Morgan Panitchpakdi, Michael J. Meehan, Kelly C. Weldon, Colette Blach, Leyla Schimmel, Rima Kaddurah-Daouk, Pieter C Dorrestein, Rob Knight, Suzanne Craft, Alzheimer’s Gut Microbiome Project Consortium

**Author notes:** Denotes equal contribution (Amanda Hazel Dilmore, Cameron Martino). Denotes corresponding authors.

## Abstract

**INTRODUCTION:** The ketogenic diet (KD) is an intriguing candidate for neuroprotection in Alzheimer’s disease (AD) given its protective effects against metabolic dysregulation and seizures. The diet’s neuroprotective effects have been shown to be gut microbiome-dependent in mice; thus we examined KD-induced changes in the gut microbiome and metabolome in patients at-risk for AD.

**METHODS:** We compared the low-carbohydrate modified Mediterranean Ketogenic Diet (MMKD) to the low-fat American Heart Association Diet (AHAD) in adults with mild cognitive impairment (MCI) and control participants. We collected stool samples for shotgun metagenomics and untargeted metabolomics at five timepoints to interrogate the microbiome and metabolome.

**RESULTS:** Individuals with MCI on the MMKD had lower levels of GABA-producing microbes *Alistipes sp. CAG:514* and GABA, and higher levels of GABA-regulating microbes *Akkermansia muciniphila*.

**DISCUSSION:** We hypothesize that MMKD protects individuals with MCI in part via influencing on GABA levels and gut-transit time.

## BACKGROUND

Despite decades of research, the pathophysiologic events that result in cognitive impairment are not fully understood, delaying the development of effective therapeutic options for neurodegenerative diseases such as Alzheimer’s disease (AD) (1, 2). However, it is now appreciated that a variety of systemic changes occur throughout the course of AD, including metabolic dysregulation (3, 4), mitochondrial dysfunction (5, 6), inflammation (7), and body compositional changes (8, 9).

AD-related metabolic dysregulation has been interrogated by comparing AD metabolomic profiles to those of unaffected individuals. However, identifying metabolites universally associated with AD or cognitive decline is not straightforward. Both the Alzheimer’s Disease Neuroimaging Initiative’s (ADNI) studies (10, 11) and mouse models (12) have demonstrated that metabolic signatures for AD are sex and *APOE* genotype-dependent. Though study of the UK Biobank cohort showed that AD incidence was correlated with metabolites including the ketone body β-hydroxybutyrate, acetone, and valine, these metabolites were also associated with age and *APOE* genotype, complicating interpretation (13). Triglycerides (particularly polyunsaturated serum triglycerides) are also broadly associated with mild cognitive impairment (MCI) and AD onset (14, 15). In terms of symptom-specific associations, both short- and medium/long-chain acylcarnitines were associated with episodic memory scores and AD severity in the ADNI cohort (16), while C3 (propionylcarnitine) was associated with decreased amyloid-β accumulation and higher memory scores (17). These observations have provided new directions for AD therapeutics.

Separately, research into the gut-microbiota brain axis has spearheaded investigation into an altered gut microbiome in AD (18). It is hypothesized that gut microbiome disruptions may lead to local gastrointestinal inflammation or disturbance of the intestinal mucosal barrier, resulting in dysregulation of the signaling pathways between the gut and brain (19). Another hypothesis for the gut microbiome in AD comes from the connection between cholesterol, bile acids, and the gut microbiota. Cholesterol metabolism is consistently reported to be dysregulated in AD and AD-related diseases (20-22). Cholesterol accumulation in the brain contributes to hepatic encephalopathy, a chronic liver disease that leads to neuron loss and increased risk for AD, through bile acid mediated effects on the farnesoid X receptor (23). This observation is significant because bile acids, the main source of cholesterol elimination in the brain, are transformed by the gut microbiota (24). One preliminary study into this link showed that bile acids in serum were associated with “A/T/N” AD biomarkers, while another demonstrated that AD-related cognitive impairment is tied to higher levels of bacterially-modified secondary bile acids and lower levels of primary bile acids (25, 26). Similarly, another group reported that among the metabolites and metabolic pathways most differential between MCI and cognitively normal (CN) groups included the deoxycholate/cholate (DCA/CA) ratio and primary bile acid synthesis (27).

A significant factor uniting the gut microbiome, inflammation, and neurocognitive status with metabolic disorders is their relationship with diet (28-31). The ketogenic diet (KD) is a candidate therapeutic for AD because of its ability to improve mitochondrial function and cerebral bioenergetics, enhance autophagy, and reduce oxidative stress (30, 31). It also reduces neuronal hyperexcitability and leads to improved amyloid and tau regulation, substantiating its potential use for cognitive impairment (32).

We previously performed a pilot trial of a low carbohydrate modified Mediterranean ketogenic diet (MMKD) in patients at-risk for AD. We found that the MMKD increased levels of Aβ42 and decreased levels of tau in the cerebral spinal fluid (CSF) as well as improved peripheral lipid and glucose metabolism (i.e. reduction of Hb1Ac, insulin, triglyceride levels) in patients with mild cognitive impairment (MCI) (32). Patients on the MMKD also showed increased cerebral ketone body uptake and cerebral perfusion, important metrics of brain function (32). Microbially, the MMKD was associated with increased relative abundances of the *Enterobacteriaceae, Akkermansia, Slackia, Christensenellaceae*, and *Erysipelotrichaceae* taxa and decreases in the relative abundances of *Bifidobacterium* and *Lachnobacterium* (33). Metabolically, the MMKD was associated with increased propionate and butyrate and decreased lactate and acetate in stool (33). Interrogation into the mycobiome, or fungal microbiome, demonstrated that MMKD increased the relative abundance of *Agaricus* and *Mrakia* and decreased the relative abundance of *Saccharomyces* and *Claviceps* in patients with MCI (34).

Although thought-provoking, previous analyses relied on amplicon sequencing (of the 16S ribosomal RNA gene and internal transcribed spacer regions). In the present study, we sought to further characterize the complex relationship between diet, cognitive status, and the human microbiome and metabolome using shotgun metagenomic sequencing and untargeted metabolomics on the same samples. Furthermore, we employed a novel reference data-driven tool to empirically and retrospectively read out dietary metabolites from our samples (35). By updating our methods and integrating multiple data types, we identified key modulations to the gut microbiome and metabolome related to diet alone, cognitive status alone, or both.

## METHODS

### Participant Information

Participants were deemed at risk for AD based on their systemic metabolic dysfunction and cognitive dysfunction or subjective/objective memory complaints at study onset. Exclusion criteria included statin use and neurological conditions other than MCI. See **Methods Supplement** for full inclusion and exclusion criteria for this study. The protocol was approved by the Wake Forest Institutional Review Board (ClinicalTrials.gov Identifier: NCT02984540), and written informed consent was obtained from all participants and/or their study partners. Participants were medically supervised by clinicians, with safety monitoring overseen by the Wake Forest Institutional Data and Safety Monitoring Committee.

### Procedure

The primary pilot trial was a randomized crossover design in which participants consumed either a Modified Mediterranean-Ketogenic Diet (MMKD) or the control American Heart Association Diet (AHAD) for 6 weeks, followed by a 6-week washout with their pre-study diet, after which the second diet was consumed for 6 weeks (**Figure S1**). For further information on diet information and education, see the **Methods Supplement**.

### Data Collection and Processing

Stool samples were collected from participants at five study timepoints: at the beginning and end of each diet intervention, and 6 weeks after washout of the second diet (**Figure S1**). Methods for collection were adapted from the Manual of Procedures for the Human Microbiome Project (NIH, Version Number 12.0). We extracted DNA for metagenomic sequencing and metabolites for untargeted MS/MS analysis. For more information on metagenomics, metabolomics, and food-omics data processing and analysis, see the **Methods Supplement**.

### Data Availability

Metagenomic sequencing data are available in Qiita under study ID 13662 (36). Mass spectrometry data (.mzXML format) are available in MassIVE under ID MSV000087087. The classical molecular networking job is available in GNPS at the following link: https://gnps.ucsd.edu/ProteoSAFe/status.jsp?task=90f1ba6e1e4d4d89b75b9017a0631983 (37). The feature-based molecular networking job is available in GNPS at the following link: https://gnps.ucsd.edu/ProteoSAFe/status.jsp?task=16e7c66221ce4ae9ae6678ec276d8343 (38). The code utilized for these analyses are available at https://github.com/ahdilmore/BEAM_MultiOmics.

## RESULTS

Twenty-three adults were enrolled in the study and twenty completed the full intervention. Participant characteristics by AD risk group may be found in **Table 1**.

**Table 1.** Participant characteristics for full sample and by AD risk group at study baseline. Table values are mean (SD) or N (%). +Denotes a difference between CN and MCI, p<0.05, using t-test for continuous and Fisher’s exact test for categorical variables. ^APOE genotype information unavailable for one participant.

| Variable | All (n=20) | CN (n=11) | MCI (n=9) |
| --- | --- | --- | --- |
| Age, years | 64.3 (6.3) | 64.9 (7.9) | 63.4 (4.0) |
| Female sex | 15 (75%) | 9 (81.8%) | 6 (66.7%) |
| Black race | 7 (35%) | 2 (18.2%) | 5 (55.6%) |
| APOE4-positive | 6 (31.6%) | 2 (20%)<sup>^</sup> | 4 (44.4%) |
| Education, years | 16.1 (2.5) | 16.5 (2.3) | 15.7 (2.9) |
| BMI, kg/m<sup>2</sup> | 28.4 (5.7) | 26.9 (6.2) | 30.3 (4.7) |
| MMSE | 28.7 (1.1) | 28.9 (1.0) | 28.3 (1.2) |
| Glucose, mg/dL | 97.6 (18.3) | 93.9 (11.4) | 102.1 (24.4) |
| BHB, mmol/L | 0.23 (0.27) | 0.35 (0.31)+ | 0.1 (0.14)+ |
| Insulin, μIU/mL | 8.3 (6.1) | 5.2 (3.4)+ | 12.0 (6.8)+ |
| HbA1c, % | 5.9 (0.3) | 5.9 (0.2) | 6.1 (0.4) |
| Total Chol, mg/dL | 215.2 (43.7) | 200.5 (42.1) | 233.2 (40.6) |
| HDL Chol, mg/dL | 67.4 (25.8) | 69.5 (25.8) | 64.8 (27.1) |
| VLDL Chol, mg/dL | 20.0 (11.1) | 15.2 (6.2)+ | 25.8 (13.2)+ |
| LDL Chol, mg/dL | 121.5 (41.2) | 104.1 (45.6)+ | 142.7 (24.2)+ |
| Triglycerides, mg/dL | 99.8 (55.7) | 75.5 (30.5)+ | 129.4 (66.4)+ |

### Dimensionality Reduction of Microbiome, Food-omics, and Metabolome datasets

To visualize relationships between samples, we performed dimensionality reduction with Robust Aitchison Principal Component Analysis (RPCA) on each -omic dataset (39; **Figure 1A**). We found that the metabolomics data separates most strongly by individual subject, though separation by Cognition Group and Dietary Sequence are also statistically significant (F=29.684, p=4.33e-6; F=23.562, p=3.25e-5; F=20.987, p=6.58e-5) while metagenomics and food-omics data separate most strongly by Cognition Group, though not significantly (F=3.896, p=0.196; F=4.114, p=0.213). However, RPCA did not account for the longitudinal nature of the dataset. Controlling for repeated measures with compositional tensor factorization (CTF) revealed consistent dietary-related patterns in PCA space regardless of diet order (40; **Figure 1B**). Results from all ANOVAs performed on CTF Aitchison beta-diversity distances from baseline are available in **Table S1**.

**Figure 1.**
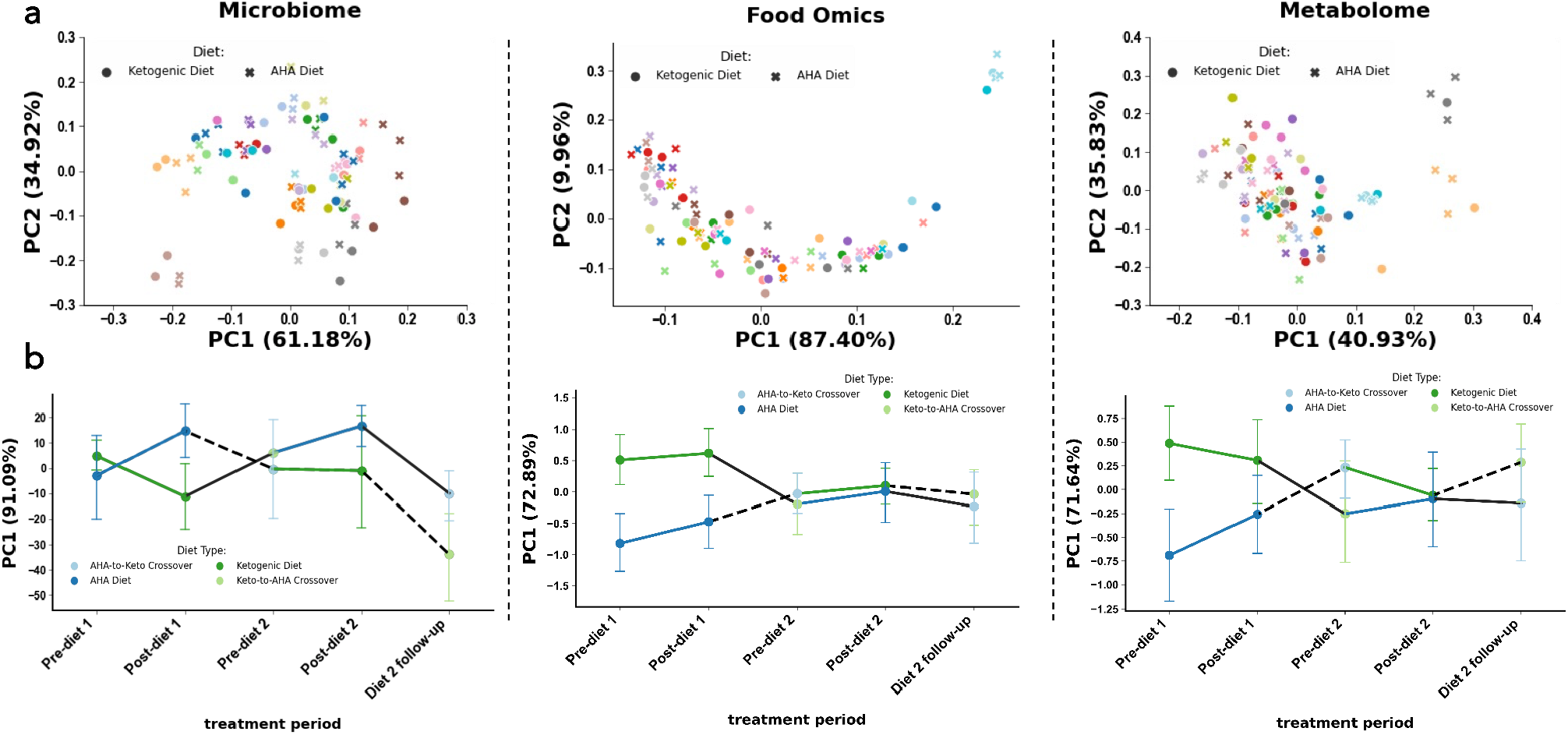
Microbiome (left column), food-omics (middle column), and metabolome (right column), separate by subject and by diet across time. Robust Aitchison Principal Component Analysis (RPCA) PC1 (x-axis) and PC2 (y-axis) colored by subject identification with marker shape by diet across study, clusters by subject. Compositional tensor factorization, method accounting from repeated measures across time (x-axis) PC1 (y-axis) colored by diet types, follows crossover study design in each data modality.

### Relative Abundance Analyses

We identified microbial, food, and metabolite features that were associated with cognition and diet using a Negative Binomial Linear Mixed Effect model. We identified features that were associated with diet but not cognitive status, cognitive status but not diet, and both diet and cognitive status.

#### Microbial Features Associated with Cognitive Status and Dietary Intervention

The top ten microbial features associated with normal cognition or mild cognitive impairment are displayed in **Figure 2A**. Notably, *Ruminococcus sp. CAG:330* is associated with MCI while *Bacteroides fragilis* and *Dialister invisus* are associated with CN independently of diet or timepoint (**Figure 2D**; p=0.016, p=0.028 respectively). The top ten microbial features associated with the ketogenic diet or a low-fat diet are displayed in **Figure 3A**. We observed higher abundances of *Akkermansia muciniphila* in the ketogenic diet, and higher abundances of *Veillonella sp. ACP1* in low-fat diets (**Figure 3D**). To identify OGUs that were highly associated with both cognitive status and diet, we plotted the log ratios for CN to MCI against the log ratios for MMKD over AHAD and examined features that fell in each corner of the plot (**Figure 4A**). We found that the ratio of *Alistipes sp. CAG:514* to *Bifidobacterium adolescentis* was significantly different between MCI and CN individuals only after consuming the low-fat AHAD (**Figure 4B**; t=3.78, p=0.00082).

**Figure 2.**
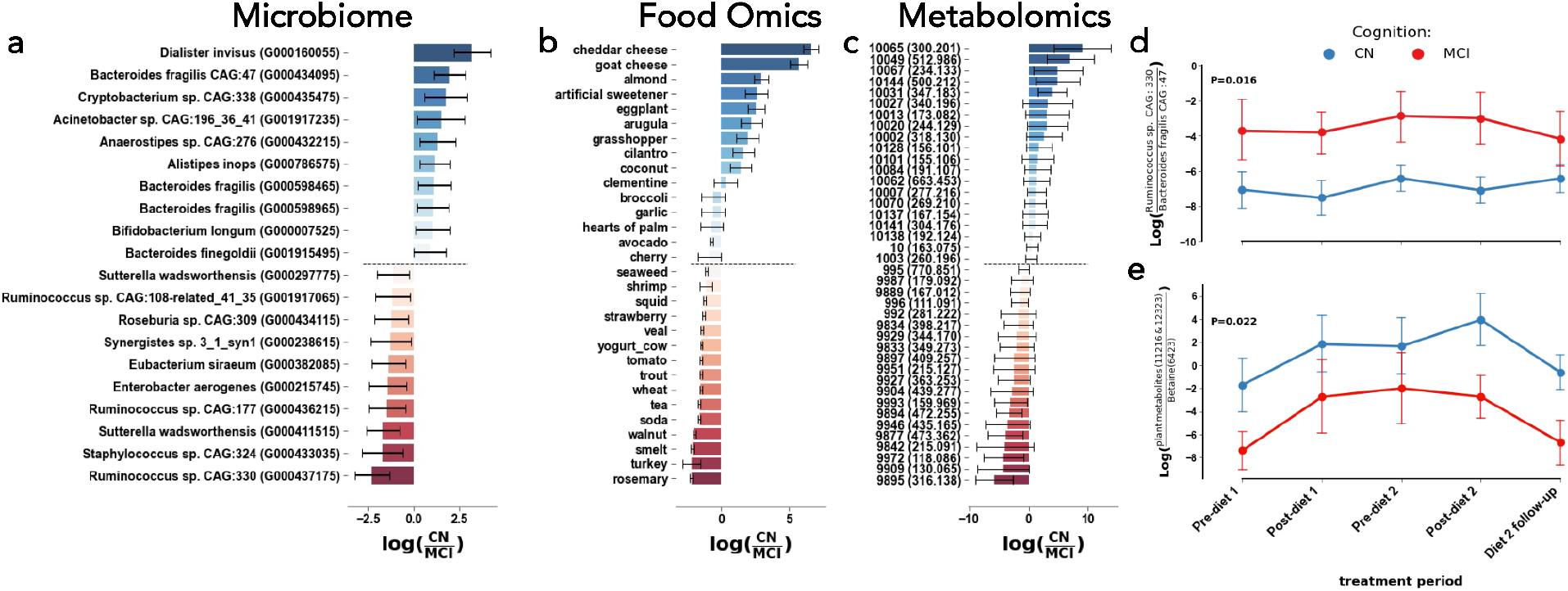
Microbial strains (left), foods (middle), and metabolites (right) related to cognition, but not necessarily diet type. The differential abundances for the top 10 and bottom 10 with mean (bar) and model 95% confidence interval (error bars) associated with cognitive normal (more positive, blue) and mild cognitive impairment (more negative, red) (**A, B, C**). The log-ratio (y-axis) of the top and bottom bacterial strains (**D&E**) plotted across time (x-axis) and colored by cognition phenotype. Presented cognition p-values in log-ratio plots are determined from linear mixed effects models with fixed effects of diet, cognition, period, and diet sequences with random effect of subject.

**Figure 3.**
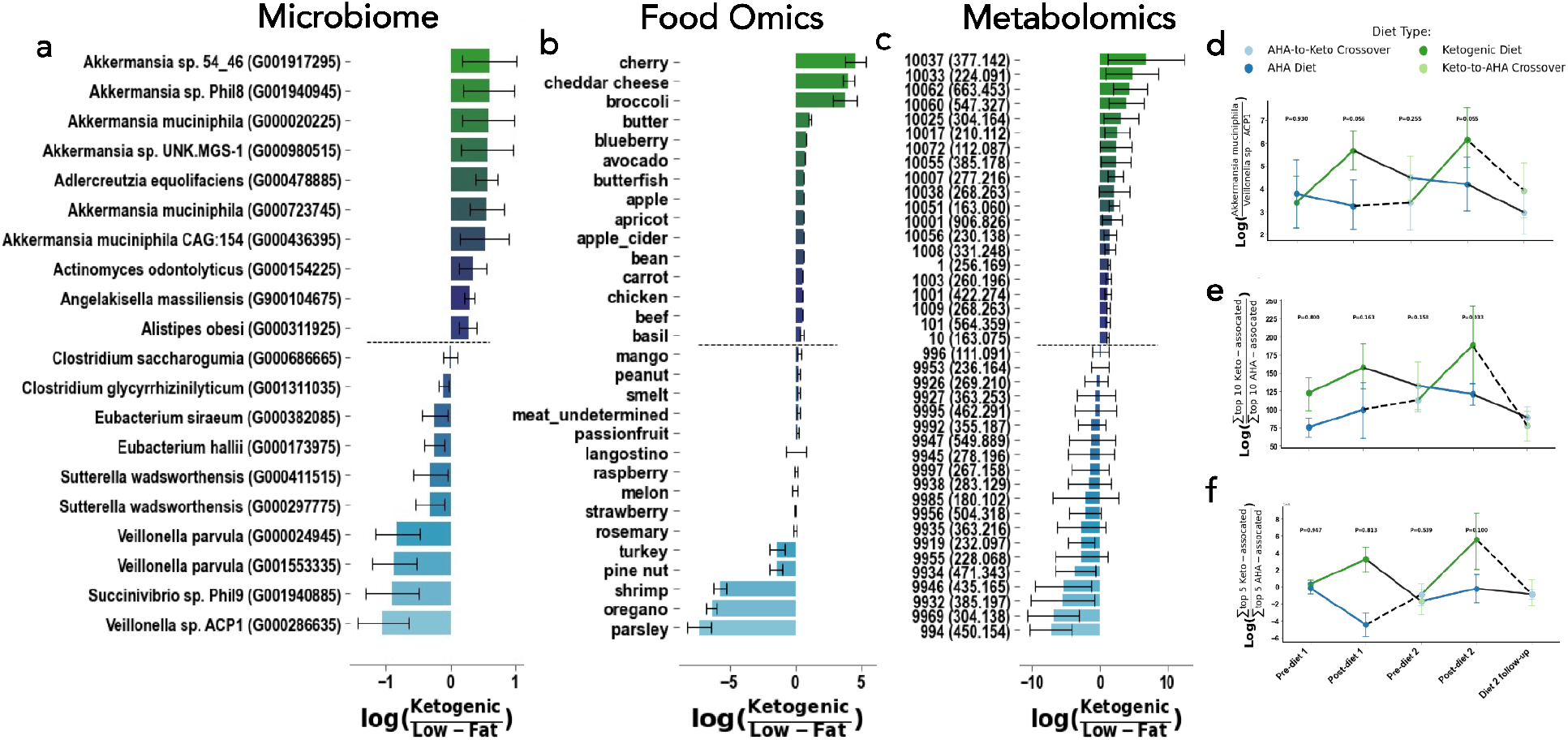
Microbial strains (left), foods (middle), and metabolites (right) related to diet type, but not necessarily cognition. The differential abundances for the top 10 and bottom 10 with mean (bar) and model 95% confidence interval (error bars) associated with cognitive Ketogenic (more positive, green) and low-fat / american heart association diets (more negative, blue) (**A, B, C**). The log-ratio (y-axis) of the top and bottom bacterial strains (**D**), and sum of the top and bottom 10 foods (**E**) or metabolites (**F**) plotted across time (x-axis) and colored by diet. Presented diet stage p-values in log-ratio plots are determined from linear mixed effects models with fixed effects of diet, cognition, period, and diet sequences with random effect of subject.

**Figure 4.**
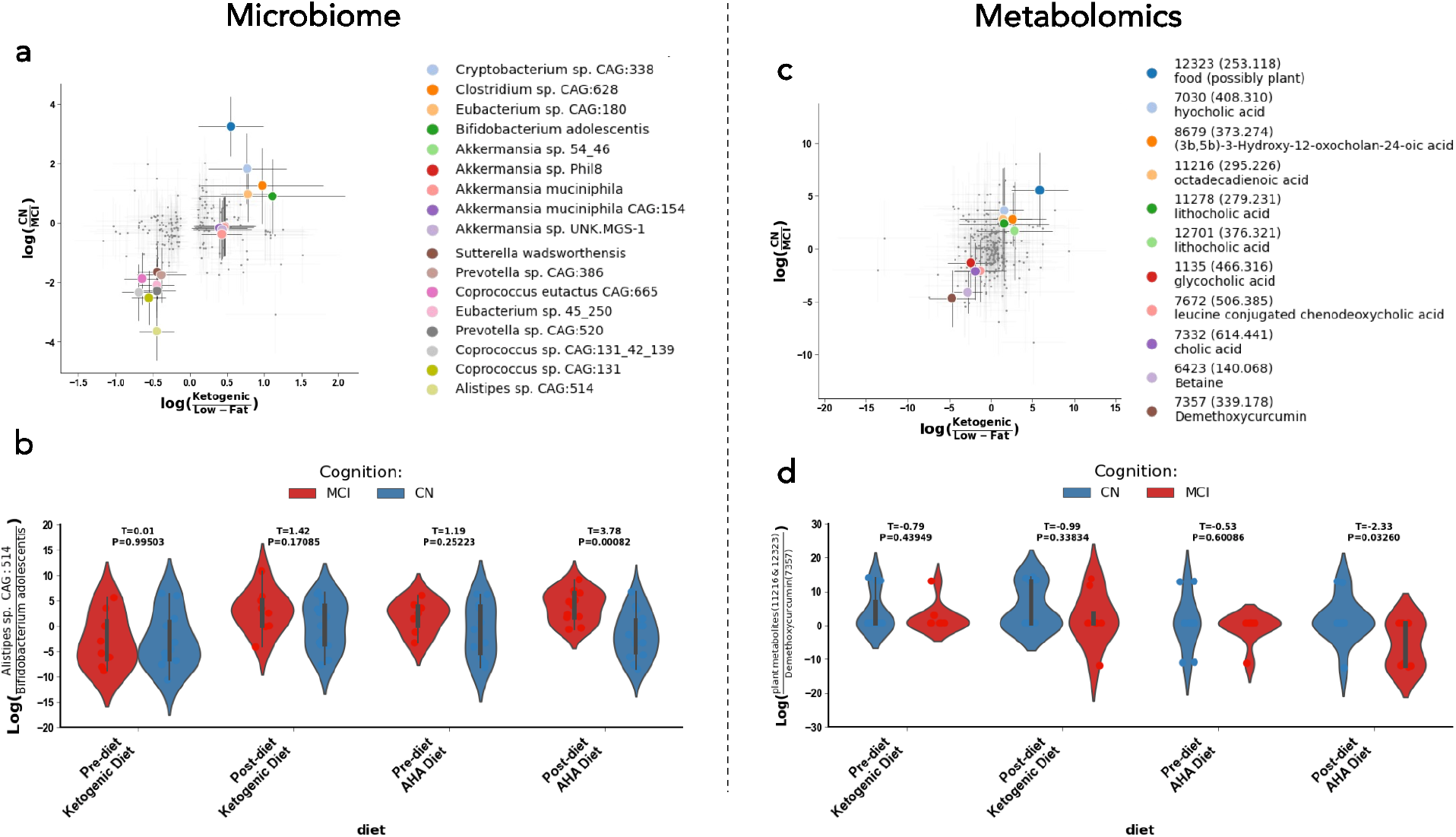
Microbial strains (left), foods (middle), and metabolites (right) related to both cognition and diet. Scatter plot of log ratios of differential abundances by diet (x-axes) and cognition (y-axes) for microbes, with the ten most differential microbes and metabolites in both axes colored (**A**). Violin plot of the log-ratio of *Alistepes sp. CAG: 514* to *Bifidobacterium adolescentis* at four different timepoints in the study (**B**). Scatter plot of log ratios of differential abundances by diet (x-axes) and cognition (y-axes) for microbes, with the ten most differential microbes and metabolites in both axes colored (**C**). Violin plot of the log-ratio of plant metabolites to Diarylheptanoids at four different timepoints in the study (**D**). In the scatterplots shown in **(A)** and **(C)**, each corner corresponds to a different subpopulation: the top right corner represents microbes/metabolites associated with the MMKD and normal cognition, the top left corner contains the microbes/metabolites correlated with the AHAD and normal cognition, the bottom left corner shows microbes/metabolites associated with the AHAD and mild cognitive impairment, and the bottom right corner corresponds to microbes/metabolites correlated with the MMKD and MCI. These analyses allowed us to examine microbial and metabolite changes that were modulated by both diet and cognitive status.

#### Food Features Associated with Cognitive Status and Dietary Intervention

The top food microbial features associated with normal cognition or MCI are displayed in **Figure 2B** and the top food microbial features associated with the MMKD or AHAD are displayed in **Figure 3B**. Interestingly, cognitively normal individuals had higher levels of dietary metabolites that matched to cheddar cheese, goat cheese, and almonds - all foods that one might consume on a ketogenic diet - while individuals with MCI had relatively higher loads of dietary metabolites that matched to turkey and smelt (**Figure 2B**). As one might expect, those on the MMKD had higher relative abundances of dietary metabolites found in dairy products including cheddar cheese, low-sugar fruits like cherries, and low-sugar vegetables including broccoli (**Figure 3B**). Those on the AHAD diet had higher relative abundances of dietary metabolites that matched to foods including parsley, oregano, and shrimp (**Figure 3B**). However, the log ratios of the sum of the top 10 most MMKD-associated to the sum of top 10 AHAD-associated dietary metabolites were not significantly different over the course of the study (**Figure 3E-F**).

#### Metabolites Associated with Cognitive Status and Dietary Intervention

The top metabolite Feature IDs associated with normal cognition or MCI are displayed in **Figure 2C**, while the top metabolite Feature IDs associated with the MMKD or AHAD are displayed in **Figure 3C**. We found that the log ratio of the sum of the top 5 metabolites associated with the ketogenic diet to the sum of the top 5 metabolites associated with the low-fat diet was not significantly different throughout the intervention (**Figure 3F**). Furthermore, none of the top metabolites mapped to an exact reference through FBMN. However, metabolite 10007, which is associated with both the ketogenic diet and cognitive normality, was suspected to be related to curcumol through a nearest neighbor suspect spectral library (41). It is interesting that curcumol is associated with the ketogenic diet and normal cognition since another *Curcuma* metabolite, curcumin, has been associated with decreased gut motility through increased both gallbladder contraction and luminal bile acid levels (42). For this reason, in tandem with the fact that cognitive impairment in AD is tied to higher levels of bacterially-modified secondary bile acids and lower levels of primary bile acids, we decided to monitor bile acid levels in this population (24). We plotted the log ratios for cognitive normal to mild cognitive impairment against the log ratios for MMKD over AHAD and examined annotated bile acids as well as other molecules related to curcumin (**Figure 4C)**. We found that the log ratio of two plant-derived metabolites to Diarylheptanoids, the class of molecules curcumin is a part of, was much lower in individuals with MCI only after consuming the AHAD (**Figure 4D**; t=-2.33, p=0.0326).This indicates that curcumin may be selectively enriched in people with cognitive impairment when they eat a low-fat diet.

### Multi-omics Analyses

We were particularly interested in monitoring the relative levels of primary and secondary bile acids in individuals with MCI and those with normal cognition due to their associations with AD. Bile salt hydrolase (BSH) promotes the production of secondary bile acids, so we sought to examine the relationship between abundance of BSH-containing microbes and bile acid levels (42, 26). We used MMvec to identify bile acids that are associated with particular microbes and to cluster these relationships by diet (43). We found that *Bifidobacterium adolescentis, Akkermansia muciniphila*, and *Akkermansia muciniphila CAG:154* were associated with 12-ketodeoxycholic acid, taurocholic acid, cholic acid, and hyocholic acid in the MMKD while *Alistipes sp. CAG:514* and *Clostridium sp. CAG:568* were associated with demethoxycurcumin, deoxycholic acid, cholic acid, and leucine-conjugated chenodeoxycholic acid in the AHAD (**Figure 5A**). We were also interested in examining the relationship between the metabolite demethoxycurcumin and levels of BSH-containing microbes as well as the ratio of unconjugated to conjugated bile acids, since turmeric increases luminal bile acid levels (42). Interestingly, we found that individuals with MCI had lower relative abundances of BSH-encoding microbes when their metabolome contained curcumin, while no difference was found in cognitively normal individuals (**Figure 5B**; t=2.9, p=0.0089; t=0.5, p=0.6308). On the other hand, cognitively normal individuals had a higher ratio of unconjugated bile acids to conjugated bile acids when their metabolome contained curcumin, while there was no significant difference in individuals with MCI (**Figure 5C**; t=-2.3, p=0.0328; t=-0.6, p=0.5681).

**Figure 5.**
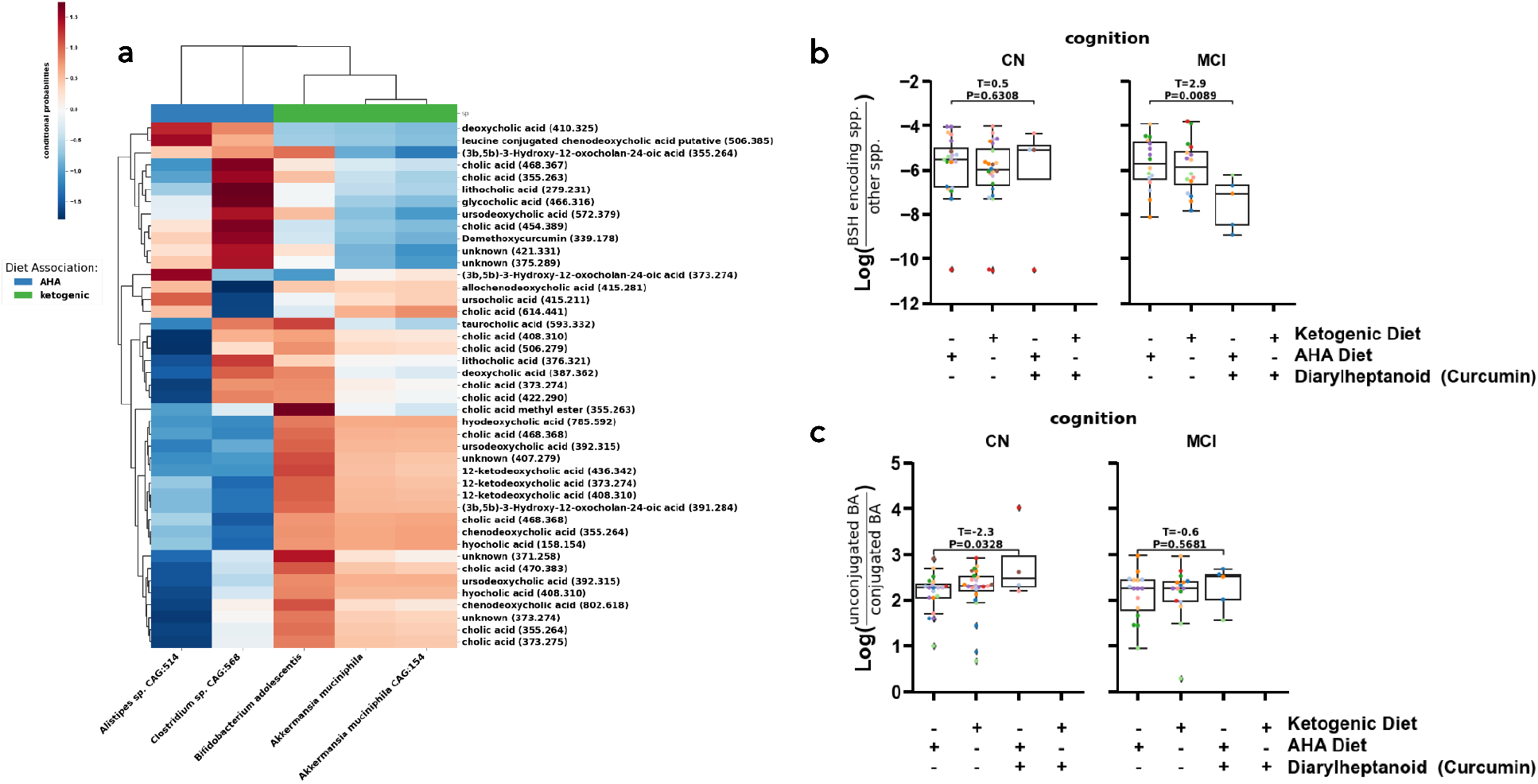
Strongest diet and cognition linked differential abundances and co-occurrences linked to bile salt hydrolase (BSH) containing bacterial species, diarylheptanoids, and bile acids (BA). The MMvec log cooccurrence probabilities (red-blue colors, colorbar) between differentially abundant and annotated metabolites (y-axis) and microbes (x-axis) colored by their association to diet (color bar top of x-axis, color legend) (**A**). The log-ratio of BSH containing microbes vs. those without BSH (y-axis) across diets and diarylheptanoids consumption (x-axis) (**B**). The log-ratio of unconjugated to conjugated BAs (y-axis) across diets and diarylheptanoids consumption (x-axis) (**C**). The annotations are Level II or Level III according to the Metabolomics Standards Initiative (46). Significance was evaluated by a two-sided t-test.

## DISCUSSION

The gut microbiome, metabolism, and neurocognition are all significantly influenced by diet. In this work, we have demonstrated the effect of two dietary interventions for mild cognitive impairment on the gut microbiome, dietary metabolites, and general metabolome.

Notably, we showed that the microbiome and food-ome were not significantly associated with cognitive status or diet without explicitly accounting for the longitudinal nature of the data. Upon doing so, we found that the metagenome, food-ome, and metabolome were significantly associated with diet. The metagenome and metabolome were not significantly impacted by dietary order, while the food-ome was. Although there was noise due to factors such as inter-individual variation, dietary sequence, and cognitive status, there were consistent diet-related variations in the microbiome. This pattern corresponds to previous observations that dietary-induced changes to the microbiome can overwhelm inter-individual differences, even when dietary changes are short-term (28).

We also identified the top microbial, metabolic, and food features that were associated with diet independent of cognition and cognition independent of diet. Microbially, we found that *Ruminococcus sp. CAG:330* is associated with MCI while *Bacteroides fragilis* and *Dialister invisus* are associated with cognitive normality. Furthermore, *Akkermansia muciniphila* is highly associated with the MMKD, while *Veillonella sp. ACP1* is related to the AHAD. The *Akkermansia muciniphila* observation matches previous reports that its relative abundance increases significantly in mice when they are fed a ketogenic diet (44). This same study demonstrated that microbe *Akkermansia muciniphila* is essential for the ketogenic diet’s ability to protect against seizures in mice since it limits gut bacterial GABA production (44). Interestingly, we found that *Alistipes sp. CAG:514*, is enriched in individuals with MCI on the low-fat AHAD (**Figure 4B**). This finding is significant because microbial strains that are similar to *Alistipes sp. CAG:514* are known to produce GABA (45). This led us to re-investigate the relative relationship between *Akkermansia muciniphila* and *Alistipes sp. CAG:514* and levels of the GABA metabolite in our population. We found that the ratio of *Alistipes sp. CAG:514* to *Akkermansia muciniphila* is elevated in individuals with MCI relative to normal individuals only on the low-fat AHAD (**Figure S3A**). Furthermore, GABA levels are higher in the stool of cognitively-impaired individuals on the AHAD than on the MMKD, but there is no dietary-related difference in cognitively-normal individuals (**Figure S3B**). This trend was consistent in a separate dataset examining cerebrospinal fluid metabolites of the same individuals (**Fig S3B**). These observations led us to hypothesize that low-fat AHAD modulates key neurotransmitters, such as promoting GABA production, in individuals with cognitive impairment. This dysregulation may be related to increases in GABA-producing microbes such as *Alistipes sp. CAG:514*, while dietary interventions such as the MMKD may decrease GABA levels in the opposite direction through its associated increases in GABA-regulating microbes such as *Akkermansia muciniphila*.

We found that one of the top metabolites associated with both the ketogenic diet and cognitive normality was related to curcumol through a nearest-neighbor suspect library (41). Another compound produced by the same plant, curcumin, was instead elevated in participants with MCI who ate the low-fat AHAD diet (**Figure 4F**). Consumption of turmeric, which contains curcumin and curcumol, is known to stimulate gallbladder contraction which results in higher luminal bile acid levels (42). Bile acids were also of interest because the cognitive impairment observed in AD is tied to higher levels of bacterially-modified secondary bile acids and lower levels of primary bile acids (24). Therefore, we hypothesized that cognitively normal people may have higher relative levels of primary bile acids when curcumin is found in their metabolome. Indeed, we found that cognitively normal individuals had a significantly higher ratio of unconjugated bile acids to conjugated bile acids when their metabolome contained curcumin, while there was no significant difference in individuals with MCI (i.e. individuals with MCI had relatively higher load of conjugated bile acids, **Figure 5C**). We hypothesized that the difference in relative levels of unconjugated to conjugated bile acids may be due to the levels of BSH (26). Cognitively normal individuals retained similar levels of BSH-containing microbes regardless of diet or curcumin-presence, while individuals with cognitive impairment had significantly lower levels of BSH-containing microbes when curcumin was present in their diet (**Figure 5B**). It appears, therefore, that the low-fat AHAD decreases the levels of BSH-containing microbes as well as *Akkermansia muciniphila* in individuals with MCI, leading to alterations of key metabolites including the neurotransmitter GABA. These individuals also have higher relative levels of unconjugated bile acids to conjugated bile acids than cognitively normal individuals, leading us to hypothesize that they may have decreased gut-transit time. We have outlined the details of our overall model in **Figure S4**.

### Limitations & Further Work

Our study is limited by a small sample size, and relatively brief intervention period. A significant methodological limitation of our study was that our untargeted metabolomics experiments left many of the key metabolites that were highly related to cognition and diet unannotated. For this reason, much of our metabolomics analyses relied on metabolites that have previously been described in the Alzheimer’s disease and dietary metabolite literature. Furthermore, since we did not run bile acid or neurotransmitter standards in these experiments, our annotations are Level II or Level III by the Metabolomics Standards Initiative (46). Since we know that bile acid metabolites and neurotransmitters such as GABA are interesting, in the future we may choose to perform targeted metabolomics or at a minimum, run these compounds as standards in the same mass spectrometry run to increase our annotation rigor.

Similarly, while our food-omics tool provides a handy reverse method to readout food counts, it is not perfect in reproducing exact dietary features. Although it reproduces dietary patterns that are consistent with the ketogenic diet or a low-fat diet, the tool cannot be used to distinguish exact dietary conditions. As more reference foods are added to the FoodOmics database and the tool grows in power, its power will grow. In the future, we may also choose to utilize patient dietary logs to identify foods that are highly associated with changes in cognition.

An additional technical limitation is that the untargeted metabolomics platform does not provide the level of characterization of lipid metabolism and mitochondrial function that is available on targeted platforms. The current results should be interpreted as a snapshot of metabolism with more detailed analysis of lipid metabolism, ketogenesis, and other processes known both to be affected by diet and important for AD to come in future studies.

Despite these technical challenges, we performed a well-controlled, longitudinal trial and used statistical techniques that controlled for various challenging aspects of our data (sparsity, compositionality, and longitudinality). These statistical analyses allowed us to identify microbial and metabolite features that were specifically associated with both diet and cognitive status.

## Supporting information

SuppFigures

SuppMethods

## Data Availability

Metagenomic sequencing data are available in Qiita under study ID 13662. Mass spectrometry data (.mzXML format) are available in MassIVE under ID MSV000087087. The classical molecular networking job is available in GNPS at the following link: https://gnps.ucsd.edu/ProteoSAFe/status.jsp?task=90f1ba6e1e4d4d89b75b9017a0631983. The feature-based molecular networking job is available in GNPS at the following link: https://gnps.ucsd.edu/ProteoSAFe/status.jsp?task=16e7c66221ce4ae9ae6678ec276d8343. The code utilized for these analyses are available at https://github.com/ahdilmore/BEAM_MultiOmics.

https://gnps.ucsd.edu/ProteoSAFe/status.jsp?task=16e7c66221ce4ae9ae6678ec276d8343

https://github.com/ahdilmore/BEAM_MultiOmics

## ACKNOWLEDGEMENTS / CONFLICTS / FUNDING SOURCES

This work was supported by the Wake Forest Alzheimer’s Disease Research Center (P30-AG072947), the Hartman Family Foundation, and the Roena B. Kulynych Center for Memory and Cognition Research.

Funding for the AGMP (Alzheimer’s Gut Microbiome Project) and ADMC (Alzheimer’s Disease Metabolomics Consortium), led by Dr R.K.-D. at Duke University) was provided by the National Institute on Aging grants 5U19AG063744, 31U01AG061359-01, 1RF1AG059093, 1RF1AG057452, and R01AG046171, a component of the Accelerating Medicines Partnership for AD (AMP-AD) Target Discovery and Preclinical Validation Project (https://www.nia.nih.gov/research/dn/amp-ad-target-discovery-and-preclinical-validation-project) and the National Institute on Aging grant RF1 AG0151550.

R.K-D. is an inventor of key patents in the field of Metabolomics and holds equity in Metabolon, a biotech company in North Carolina. In addition, she holds patents licensed to Chymia LLC and PsyProtix with royalties and ownership, which is unrelated to this work. P.C.D. is on the scientific advisory board of Sirenas, Cybele Microbiome, Galileo, and founder and scientific advisor of Ometa Labs LLC and Enveda (with approval from UC San Diego).

## Notes

### Clinical Trial

NCT02984540

### Author Declarations

The protocol was approved by the Wake Forest Institutional Review Board (ClinicalTrials.gov Identifier: NCT02984540), and written informed consent was obtained from all participants and/or their study partners. Participants were medically supervised by clinicians, with safety monitoring overseen by the Wake Forest Institutional Data and Safety Monitoring Committee.

