## Supplementary material for "Effects of a Ketogenic and Low Fat Diet on the Human Metabolome, Microbiome and Food-ome in Adults at Risk for Alzheimer’s Disease": SuppFigures

**SUPPLEMENTARY FIGURES**

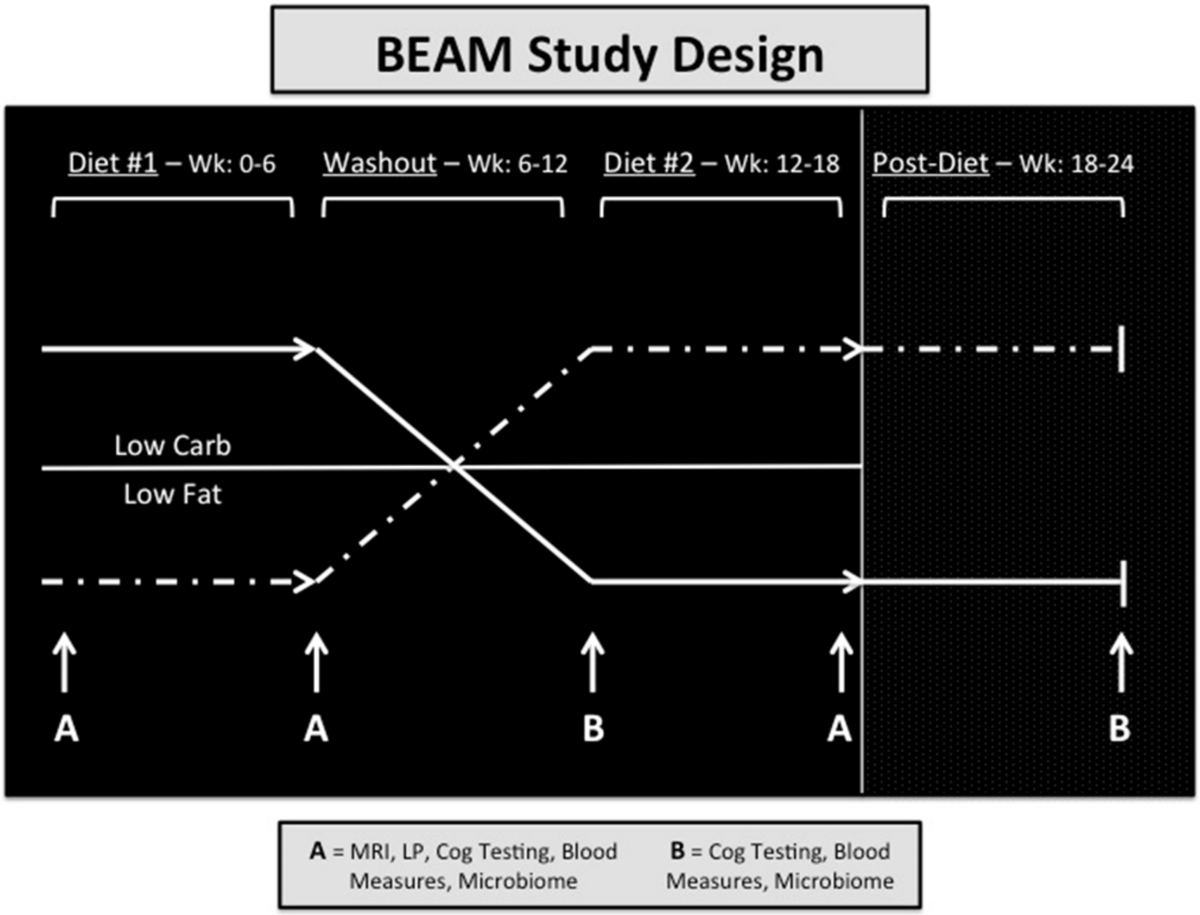


**Figure S1. BEAM Study design.** The primary study was a randomized cross-over design of two dietary interventions: the high-fat Modified Mediterranean Ketogenic Diet and low-fat American Heart Association Diet. With the cross-over design, each participant acts as their own control to assess dietary impact on study measures (i.e. microbiome, metabolome, food-ome), which were collected at five timepoints. See Neth et al. for complete details and results from the primary study (32).

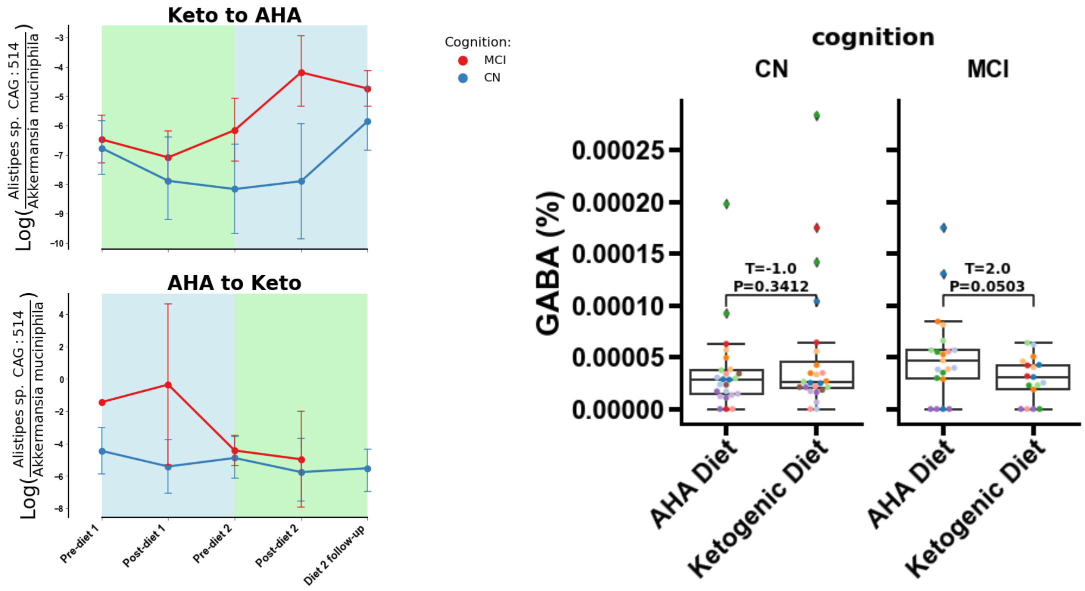


**Figure S2. Levels of GABA-producing *Alistepes* and GABA in stool.** Scatter plot of the log ratio of *Alistepes sp. CAG: 514* to *Akkermansia muciniphila* in MCI and CN individuals who started on the MMKD and transitioned to the AHAD (**A**) and individuals who started on the AHAD and transitioned to the MMKD (**B**). Boxplot showing the relative abundance of the GABA metabolite in stool samples collected from CN and MCI individuals on the AHAD and the MMKD (**C**). Significance was evaluated with a two-sided t-test.

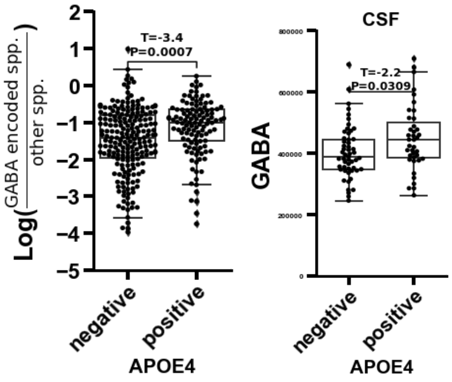


**Figure S3. Levels of GABA-producing species in stool and GABA in cerebrospinal fluid (CSF).** Scatter plot of the log ratio of GABA-encoding species to all other species in the stool of individuals who are APOE4-negative or APOE4-positive (**A**). Scatter plot of the relative abundance of GABA in the CSF of individuals who are APOE4-negative or APOE4-positive (**B**). Significance was evaluated with a two-sided t-test.

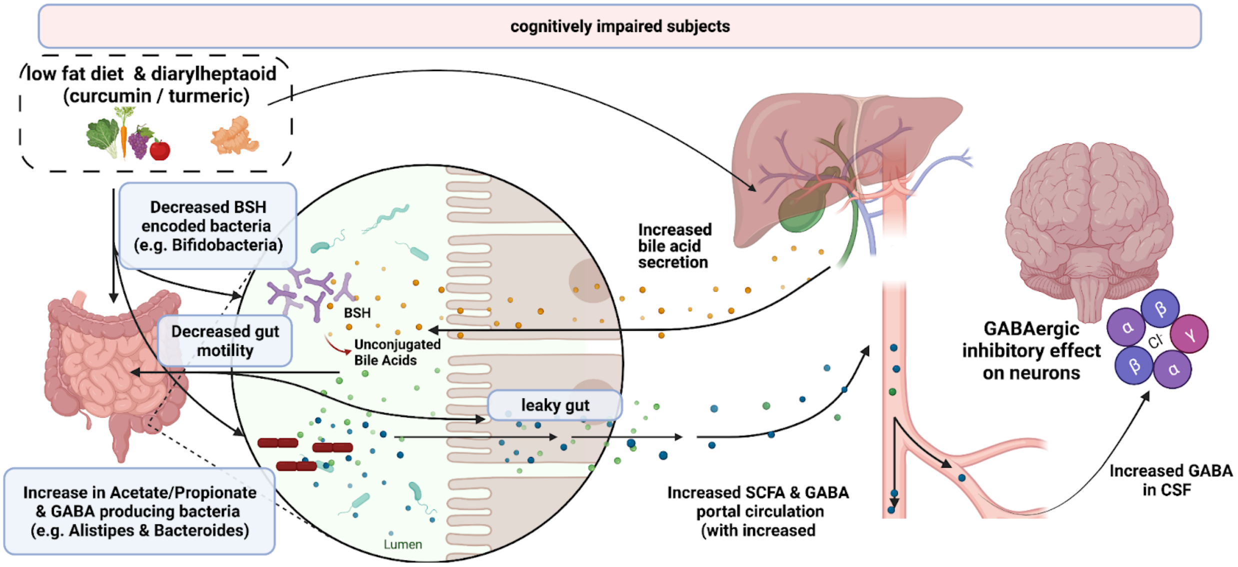


**Figure S4. The AHAD modulates key microbes and metabolites that decrease gut motility in individuals with mild cognitive impairment.** Based on our results, we propose a model for decreased gut motility and increased GABA levels in the stool and cerebrospinal fluid of individuals with mild cognitive impairment. The low fat diet, in conjunction with consumption of dietary curcumin, leads to an increase in short chain fatty acids (acetate, propionate) and GABA through an increase in bacteria that produce these metabolites (such as *Alistipes* and *Bacteroides*). There is also a decrease in BSH-encoding bacteria, such as *Bifidobacteria*. This leads to leaky gut symptoms and decreased gut motility, and promotes higher GABA levels in CSF, which can lead to inhibitory effects on neurons.
