## Supplementary material for "Effects of a Ketogenic and Low Fat Diet on the Human Metabolome, Microbiome and Food-ome in Adults at Risk for Alzheimer’s Disease": SuppMethods

**METHODS SUPPLEMENT**

**Participant Information:**
All participants were deemed to be at risk for AD based on their systemic metabolic dysfunction and cognitive dysfunction or subjective memory complaints at study onset (see **Table 1** for full participant characteristics). Criteria for metabolic dysfunction was a hemoglobin A1c of level 5.7-6.4%, which corresponds to the American Diabetes Association pre-diabetes category according to their 2016 guidelines (1). Subjective memory complaints (SMC) were diagnosed using the Alzheimer's Disease Neuroimaging Initiative (ADNI) criteria and mild cognitive impairment (MCI) was diagnosed by expert physicians and neuropsychologists using NIA-AA guidelines for clinical diagnoses without reference to biomarker status (2, 3). Study exclusion criteria included prior diagnosis of neurological or neurodegenerative illness (except MCI), major psychiatric disorder (although well-controlled depression was allowed), prior stroke, current use of diabetes and lipid lowering medications, or medications with known effects on the central nervous system (i.e. anti-seizure medications, anti-psychotics, opioids, etc.).

The protocol was approved by the Wake Forest Institutional Review Board (ClinicalTrials.gov Identifier: NCT02984540), and written informed consent was obtained from all participants and/or their study partners. Participants were medically supervised by clinicians, with safety monitoring overseen by the Wake Forest Institutional Data and Safety Monitoring Committee.

**Procedure:**
The primary pilot trial was a randomized crossover design in which participants consumed either a Modified Mediterranean-Ketogenic Diet (MMKD) or the control American Heart Association Diet (AHAD) for 6 weeks, followed by a 6-week washout, after which the second diet was consumed for 6 weeks (**Figure S1**). During the washout period participants were instructed to resume their pre-study diet and specifically not continue the protocols from their first diet. Randomization was performed with a random number generator so that the number of MCI and SMC participants in each dietary group would be roughly equal. Baseline cognitive, imaging, blood, stool, and CSF measures were obtained prior to diet randomization (4).

**Diet Intervention and Education:**
The experimental Modified Mediterranean Ketogenic Diet (MMKD) was a very low carbohydrate diet aimed at inducing ketosis. Notably, modified versions of the KD have increasingly been utilized in cases of medically intractable epilepsy given its similar efficacy and increased tolerability for patients (5). These modified KDs generally incorporate slightly higher amounts of carbohydrates from whole food sources, yet are still very low carbohydrate and reliably induce ketosis. The control diet was adapted from the low-fat American Heart Association Diet (AHAD, 5-6). The diets were customized to each participant's baseline caloric needs to maintain their current weight throughout the course of the study. Participants were also asked to keep their exercise and physical activity level stable throughout the study.

The proportions of carbohydrates and fat were the main variables manipulated between the two diets. The target macronutrient composition (expressed as % of total calories) was 5-10% carbohydrate, 60-65% fat, and 30% protein for the MMKD and 55-65% carbohydrate, 15-20% fat, and 20-30% protein for the AHAD. Participants on the MMKD were asked to keep their daily carbohydrate consumption below 20g each day throughout the 6-week intervention, while higher fat foods (preferably those low in saturated fats) could be consumed liberally. They were encouraged to eat fish, lean meats, leafy green vegetables, and nuts and discouraged from consuming artificially sweetened beverages or products marketed as “low-carbohydrate” during the intervention. Participants on the AHAD were encouraged to limit their amount of fat intake to 40g or less each day, and to eat plentiful fruits, vegetables, and fiber-laden carbohydrates.

A registered dietitian developed daily meal plans for each participant based upon their food preferences and caloric needs as determined by a pre-study 3-day food diary, body composition, and activity level. Participants had weekly diet education visits (either in-person or by phone) starting one week prior to the start of each diet and continuing throughout the remainder of the intervention. Participants maintained a daily food record that was reviewed at these visits.

Adherence to the dietary intervention was assessed by capillary ketone body (beta-hydroxybutyrate) measures that were collected at all major time points and during diet education visits using the Nova Max Plus® (http://www.novacares.com/nova-max-plus/) and with participant subjective report recorded by study dietician. Prior evidence suggests that less frequent blood-based ketone body measures are equally accurate as daily urine ketone body test strips in assessment of ketosis (7).

For the most part, participants were required to supply their own food. They were given a food stipend of $25 each week provided to offset the higher food costs associated with switching their diets. All participants were provided with a daily multivitamin supplement (Centrum® Silver®), and participants on the MMKD were supplied with 1L bottles of extra virgin olive oil during their Pre-Diet and Mid-Diet visits to promote consumption of healthy fats. Participants were not allowed to use resveratrol, CoQ10 (coenzyme Q10), curcumin, coconut oil or other medium chain triglyceride-containing supplements throughout the duration of the study.

**Stool Collection**

Stool samples were collected from participants at five study timepoints (**Figure S1**). Methods for collection were adapted from the Manual of Procedures for the Human Microbiome Project (NIH, Version Number 12.0). Participants received gloves, a sterile collection container, sterile scooping tool, and toilet hat in addition to instructions for proper specimen collection and storage (refrigeration or on ice) prior to transfer to study staff. Once received, samples were immediately aliquoted into four individual vials from each timepoint and stored at -80°C until further processing and analyses.

**Metagenomic Sample Processing:**

DNA was extracted from stool samples according to Earth Microbiome Project protocols (8) using the QIAGEN® MagAttract® PowerSoil® DNA KF Kit (384-sample). A total of 5 ng (or 3.5 μL maximum) genomic DNA were used in a 1:10 miniaturized Kapa HyperPlus protocol with a 15-cycle PCR amplification for shotgun metagenomic sequencing. Libraries were quantified with the PicoGreen dsDNA assay kit, and 50 ng (or 1 μL maximum) of each library was pooled. The pool was size selected for 300 to 700 bp and sequenced as a paired-end 150-cycle run on an Illumina HiSeq 4000 sequencer at the UCSD IGM Genomics Center (9).

**Metagenomic Data Processing:**

Shotgun sequencing data were uploaded to and processed by Qiita (10, Study ID 13662). Human reads were removed using minimap2 2.17 (11), while adapters, quality filtering, and trimming were performed using fastp 20.1 (12). Remaining reads were recruited to the Web of Life database (13) with Bowtie2 v2.3.0 (14) using the parameters from the SHOGUN pipeline (15), then processed into Operational Genomic Units with Woltka (16). The resulting feature table was used for downstream analysis.

**Metabolomic Sample Processing:**

To extract metabolites from fecal samples, frozen samples were thawed on ice for 30 minutes. Then, a solution of 50% methanol spiked with 1 μM sulfamethazine was added to each fecal sample (approximately 50-100 mg feces) at a volume ratio of 10 μL extraction solvent to 1 mg sample. Samples were homogenized at 25 Hz for 5 minutes on a tissue homogenizer, then centrifuged at maximum speed for 15 minutes at 4°C. A 200 μl aliquot of supernatant from each sample was transferred into a 96-well plate and vacuum concentrated to dryness via centrifugal lyophilization (Labconco Centrivap). Once dried, the samples were stored at -80 °C until LC-MS was performed. Untargeted LC-MS was performed using a Vanquish liquid chromatography system (Thermo Scientific) paired with QExactive mass spectrometer. Samples were separate using a C18 column (Phenomenex Kinetex 1.7 μm C18 100 Å LC Column 50 x 2.1 mm). The mobile phase used was LC-MS grade water (phase A) and LC-MS grade acetonitrile (phase B), both containing 0.1% formic acid (Fisher Scientific, Optima LC-MS). The LC was programmed with a flow rate set to 0.5 mL/min. Samples were injected at 95%A:5%B, which was held for 1 minute, before ramping up to 100%B over 7 minutes, which was held for 0.5 minutes before returning to starting conditions. The orbitrap mass spectrometer was equipped with a HESI-II ESI probe. The mass spectrometer was programmed to use a data-dependent acquisition method that acquired MS full scan spectra, followed by MS/MS spectra of the top 5 most abundant ions. Precursor ions were fragmented once before being added to an exclusion list for 30 seconds. Data were collected in positive ion mode.

**Metabolomics Data Processing:**

Raw Q Exactive files were converted to .mzXML format using the ProteoWizard tool MSConvert (17) software, then these reformatted files were imported to MZmine (version 2.53; 18). We performed feature finding using the parameters recommended for Feature Based Molecular Networking (FBMN) in the Global Natural Product Social Molecular Networking (GNPS) ecosystem in its documentation (19, 20). Only peaks that had an MS2 scan were retained. We exported the feature quantification table (.csv) and MS/MS spectral summary (.mgf) from

MZmine into GNPS and performed FBMN using release 28.2 with the default parameters (19, 20)

**Food-omics Data Processing:**
Food counts were inferred from metabolomics data using the "Global FoodOmics" project (http://www.globalfoodomics.org) reference data set. This dataset contains 3,579 food and beverage samples contributed by community members, that were systematically annotated (21). Matching up metabolite spectra from stool samples to metabolites identified in reference food samples enables estimation of foods consumed without the need for food frequency questionnaires (22). Specifically, each "food count" corresponded to an individual consensus node in the molecular networking results that matched to a node found in reference food sample molecular networks. Consensus nodes were required to match the specific sample type, GFOP, and not match to any of the other experiment groups. Infrequent food types that occurred less often than water (which is considered blank) were removed to filter out sporadic random matches.

**Dimensionality Reduction Analysis:**
Initial dimensionality reduction analysis was performed by Robust Aitchison Principal Component Analysis (RPCA, 23) through the gemelli plugin to QIIME2 (24; https://github.com/biocore/gemelli). Dimensionality reduction that accounted for the repeated measures study design was performed with compositional tensor factorization (CTF, 25), also through the gemelli plugin for QIIME2. The biplots output from these methods were visualized with EMPeror (26; https://github.com/biocore/emperor). The statistical significance of results were evaluated with ANOVAs comparing beta-diversity distances to baseline (27). Multiple test correction was performed through Bonferroni correction.

**Relative Abundance of Features Analysis:**
We identified metagenomic, food-omics, and metabolomic features that are associated with diet and cognition by performing Bayesian inferential regression (https://github.com/gibsramen/BIRDMAn). We utilized a Negative Binomial Linear Mixed Effect model to ensure that our statistics were in agreement with our study design; we modeled time, dietary sequence (whether MMKD intervention was first or second), cognitive status, and diet (whether a given individual was on MMKD or AHAD at a given time point) as fixed effects while subject identity was modeled as a random effect. Microbes, food features, and metabolites were ordered by the log ratio of their relative abundances in objectively normal cognition to mild cognitive impairment or MMKD to AHAD individuals, respectively (28) and the top and bottom ten features were examined more closely.

**Multi-omics Analysis:**
Microbe and metabolite co-occurrence probabilities were calculated with MMvec's paired-omics function (29; https://github.com/biocore/mmvec). Heatmap visualizations were also made with MMvec. The metabolic capabilities of the microbes were evaluated with MetaCyc (30) on the Web of Life genomes (13). Any genomes containing BSH (E.C. 3.5.1.24) were considered BSH-containing/encoding.

**Data Availability:**

Metagenomic sequencing data are available in Qiita (10) under study ID 13662. Mass spectrometry data (.mzXML format) are available in MassIVE under ID MSV000087087. The classical molecular networking job is available in GNPS at the following link: https://gnps.ucsd.edu/ProteoSAFe/status.jsp?task=90f1ba6e1e4d4d89b75b9017a0631983. The feature-based molecular networking job is available in GNPS at the following link: https://gnps.ucsd.edu/ProteoSAFe/status.jsp?task=16e7c66221ce4ae9ae6678ec276d8343.
The code utilized for these analyses are available at https://github.com/ahdilmore/BEAM_MultiOmics.
